# MeshScope-Region: Distribution, Road-Network Accessibility, and Nine-Year Evolution of ICU and HCU Capacity Across Japan’s 330 Secondary Medical Areas

**DOI:** 10.64898/2026.07.17.26358374

**Authors:** Kunihisa Ohno, Masato Hirai, Satoru Hashimoto

## Abstract

**Background:** In Japan, health planning is organized around secondary medical areas (SMAs; niji-iryo-ken; 330 areas in the 2025 classification), yet nationwide analyses of intensive care unit (ICU) capacity have been conducted mainly at the prefecture level, and a recent SMA-level study addressed only the presence or absence of ICUs. The full supply structure of intensive and intermediate critical care — ICU and high care unit (HCU) beds — has not been characterized at the SMA level with respect to its composition, road-network accessibility, and evolution over time.

**Methods:** We developed MeshScope-Region, an analytical platform built on the Hospital Bed Function Reports (byosho-kino-hokoku) for fiscal years 2016–2024, in which ICU and HCU beds were identified from notified reimbursement categories and aggregated to SMAs. Three analytical layers were integrated: (1) cross-sectional distribution of ICU/HCU beds; (2) nationwide road-network accessibility computed with the Open Source Routing Machine (OSRM) from 176,962 populated 1-km census grid cells to all facilities reporting ICU or HCU beds; and (3) a nine-year longitudinal analysis of supply-structure types, classified by k-means (k = 6) in an 8-dimensional PCA space anchored to fiscal year 2024, with earlier years projected into the same space.

**Results:** In fiscal year 2024, 20,631 ICU/HCU beds were reported nationally (7,114 ICU-type; 13,517 HCU-type) at 1,044 facilities. Zone-level totals among SMAs with any beds ranged 229-fold (3–688 beds); the 90th/10th percentile ratio of per-capita density was 3.6. In total, 90.1% of the population resided within 30 minutes’ drive of a facility with ICU beds and 97.8% within 60 minutes; only 0.8% resided beyond 90 minutes. Although 140 of the 330 SMAs had no ICU facility within their own boundaries, 84.7% of their residents could reach an ICU facility in an adjacent area within 60 minutes’ drive. Longitudinally, supply structures were highly persistent: 63.0% of SMAs (208/330) retained the same structural type across all nine years, adjacent-year rank correlations of a supply-vulnerability index were 0.887–0.924 (2016 vs. 2024: ρ = 0.711), and the number of SMAs with zero ICU beds remained frozen at 133– 141. The Gini coefficient of bed distribution declined from 0.384 to 0.262 — although computed on ICU-type beds alone it remained 0.365 in fiscal year 2024 — and capacity growth (total +27.9%) was driven predominantly by HCU beds (+41.6%) while ICU beds grew only +8.0%.

**Conclusions:** Japan’s critical care supply structure is regionally rigid, with a stable set of approximately 140 SMAs lacking ICU beds for nearly a decade, yet road-network accessibility substantially mitigates the consequences of zone-level absence. Recent capacity growth — and much of the apparent equalization — has occurred predominantly in intermediate care. MeshScope-Region provides a standing, reproducible evidence base at the geographic unit of Japan’s medical planning cycles.

## 1. Introduction

The COVID-19 pandemic demonstrated that the geographic distribution of critical care capacity, not merely its national aggregate, determines whether health systems can absorb surges in severe illness. In Japan, intensive care units (ICUs) and high care units (HCUs) — the latter an intermediate care tier with a 1:4 or 1:5 nurse-to-patient ratio — together constitute this capacity, and both are defined administratively through notified reimbursement categories.

Japanese health planning is organized around secondary medical areas (SMAs; niji-iryo-ken), the statutory geographic units within which prefectures plan hospital services under the Medical Care Act. Prefectural medical plans, revised on a six-year cycle (currently the 8th Medical Plan, 2024–2029), allocate resources and define referral structures at this granularity. The number of SMAs has itself contracted through successive consolidations — from 344 in fiscal 2016–2017 to 335 in 2018–2023 and 330 in the current (2025) classification — so that longitudinal analysis requires a consistent zone crosswalk. Rational planning therefore requires SMA-level evidence on where critical care beds exist, how accessible they are by road, and how the supply structure has evolved.

Nationwide evidence has, however, been concentrated at the prefecture level. Ohbe and colleagues documented an 8.6-fold variation in ICU beds per 100,000 population across the 47 prefectures and its association with the incidence and mortality of mechanical ventilation [1], and related work has characterized intermediate care units and their outcomes [2]. SMA-level compilations by the Japan Medical Association Research Institute cover physicians and general beds but not ICU/HCU-specific categories [4], the Japanese Society of Intensive Care Medicine has published prefecture-level tabulations of ICU and HDU beds [5], and geographic accessibility analyses of emergency care have been regional in scope [6]. At the level of individual facilities, Matsuda has proposed typologies of hospital and ward function using the same publicly released Hospital Bed Function Report data together with DPC data [8]; the unit of analysis in that line of work, however, is the hospital rather than the planning area. Most recently, Ohbe and colleagues described, for the first time at the SMA level, the national distribution of areas without ICUs and the outcomes of their residents, using the Diagnosis Procedure Combination database linked to Hospital Bed Function Reports [3].

That study established that SMA-level absence of ICUs is common and consequential. It did not, however, address three questions that planning at this level requires: the composition of the full ICU/HCU supply structure (rather than the binary presence of ICUs), the road-network accessibility of critical care for residents of areas without it, and the stability of these structures over time. We therefore developed MeshScope-Region, a platform integrating (i) SMA-level mapping of ICU and HCU beds from nine years of Hospital Bed Function Reports, (ii) an embedding-based structural typology of supply [7], (iii) nationwide drive-time accessibility computed from populated 1-km census grid cells to every facility reporting ICU or HCU beds, and (iv) a nine-year analysis of structural persistence. To the best of our knowledge, no previous study has combined these elements at the geographic unit at which Japanese medical plans are actually written.

## 2. Methods

### 2.1 Data sources

The primary data source was the Hospital Bed Function Report (byosho-kino-hokoku), an annual statutory reporting system in which all hospitals with general or long-term care beds report ward-level bed counts and notified reimbursement categories to the Ministry of Health, Labour and Welfare (MHLW). Reports for fiscal years 2016 through 2024 were used. Beds were classified as ICU-type when the ward held a notification for specific intensive care management fee categories 1–6 (tokutei-shuchu-chiryoshitsu kanri-ryo), emergency life-saving admission fee categories 2 or 4 (kyumei-kyukyu nyuin-ryo; staffed at ICU level), or the pediatric intensive care management fee; and as HCU-type for high care unit management fee categories 1–2, emergency life-saving admission fee categories 1 or 3, or the stroke care unit management fee. This assignment of emergency life-saving categories follows the convention of the Japanese Society of Intensive Care Medicine [5]. Following the fiscal year 2026 revision of the reimbursement system (effective June 2026), these categories were consolidated — specific intensive care management fee grades 1–6 into grades 1–3, and emergency life-saving admission fee grades 2/4 and 1/3 into grades 1 and 2, respectively; all data in this study precede this revision and are classified under the pre-revision scheme. Grades 5 and 6 of the specific intensive care management fee were themselves introduced in the fiscal 2024 revision and are absent from earlier report years. Neonatal and perinatal intensive care categories (NICU, MFICU, and related) were excluded. The taxonomy was cross-checked against the facility-standard notification lists published by the MHLW regional bureaus. Facilities were geocoded from reported addresses using the geocoding service of the Geospatial Information Authority of Japan, with manual verification of ambiguous addresses, and assigned to SMAs using the 330-zone classification in force in 2025, via a validated crosswalk covering the consolidations from the earlier 344- and 335-zone classifications; zone codes missing in some report years (part of fiscal 2017 and fiscal 2018–2021) were imputed from the most recent observed value for the facility. Coverage of the current classification was complete: all 330 SMAs had analyzable reports in every panel year. Resident population for the structural typology was fixed at 2025 municipal estimates so that observed changes reflect the supply side only; the accessibility analysis used the 2020 Population Census 1-km grid population (e-Stat, T001100).

### 2.2 Data integration pipeline

Report files were normalized and loaded into a cloud data warehouse (Google BigQuery) organized in a medallion architecture, from which facility-level and SMA-level aggregates were derived. Facility names and addresses were normalized and records deduplicated across reporting years. A zone-code crosswalk was constructed and validated so that all years are expressed in the 2025 SMA classification. To ensure reproducibility, the integrated facility table and crosswalk were frozen as a versioned snapshot (July 2026), on which all analyses reported here were performed.

### 2.3 Cross-sectional distribution

For fiscal year 2024, we computed SMA-level totals of ICU-type and HCU-type beds, beds per 100,000 population, and facility counts, and summarized inequality by the range across zones, the 90th/10th percentile ratio of per-capita density among zones with any beds, and the Gini coefficient. Choropleth maps were generated at the SMA level.

### 2.4 Road-network accessibility

Drive times were computed with OSRM on the OpenStreetMap road network for Japan (retrieved July 10, 2026). Origins were the 176,962 populated 1-km census grid cells, covering 126,035,175 routed residents (1,432,234 residents could not be assigned to an SMA and were excluded). For each grid cell, drive times were computed to the nearest of the 1,044 facilities reporting ICU beds (or ICU/HCU beds, respectively), using a k-nearest prefilter (k = 60) followed by exact routing. Population coverage was aggregated in 30-, 60-, and 90-minute bands nationally and, by assigning each grid cell to its SMA, per SMA in the 330-zone classification. In-zone ICU presence was defined from the same bed reports (at least one facility with ICU-type beds in the zone), so that the accessibility and capacity layers share a single definition. Two remote island SMAs could not be road-routed and are reported separately.

### 2.5 Structural typology and nine-year longitudinal analysis

Each SMA-year was represented by a standardized feature vector describing its critical care supply (ICU beds, HCU beds, per-capita densities, facility counts, and composition) and classified into one of six structural types by k-means clustering. To make types comparable across years, the standardization parameters, an 8-dimensional PCA basis, and the k-means centroids (k = 6) were fitted on fiscal year 2024 and fixed; earlier years were projected into this anchor space and classified with the same centroids, avoiding the label-correspondence problem of year-by-year re-clustering. The embedding methodology is described in detail in a companion manuscript in preparation [7]; the procedure as applied here is fully specified by the standardization, PCA basis, and centroid parameters stated above.

Structural persistence was quantified on the complete panel of 330 SMAs observed in all nine years as (i) the proportion of SMAs retaining a single structural type; (ii) the distribution of the number of type changes; and (iii) a 2016→2024 transition matrix. In addition, a supply-vulnerability index was computed for each SMA-year as the within-year rank average of the Mahalanobis distance and the k-nearest-neighbor distance (k = 8) to the anchor cloud, and its temporal stability was assessed by Spearman rank correlations between all pairs of years. Aggregate trends were summarized by the annual Gini coefficient of bed distribution, the count of SMAs with zero ICU beds, and national ICU-type and HCU-type bed totals.

### 2.6 Platform implementation

MeshScope-Region is implemented as a web application (Node.js) deployed on Google Cloud Run, serving interactive SMA choropleths for each fiscal year, per-zone structural-type history strips, accessibility layers, and national trend panels from precomputed JSON payloads. The analytical platform is available from the corresponding author upon reasonable request.

### 2.7 Ethics

This study used only publicly available administrative data describing medical institutions; no patient-level or otherwise personal data were used. Ethical review was therefore not required under the Japanese Ethical Guidelines for Medical and Biological Research Involving Human Subjects.

## 3. Results

### 3.1 National capacity in fiscal year 2024

In fiscal year 2024, 20,631 ICU/HCU beds were reported nationally: 7,114 ICU-type and 13,517 HCU-type, corresponding to 1,781 facility-category notifications at 1,044 unique facilities. All 330 SMAs of the 2025 classification were covered. Of these, 74 SMAs reported no ICU or HCU beds of any kind, and 140 reported no ICU-type beds. Zone-level totals among SMAs with any beds ranged from 3 to 688 (a 229-fold difference), while the median SMA density was 12.6 beds per 100,000 population. Population adjustment narrowed but did not eliminate the disparity: the 90th/10th percentile ratio of per-capita density among zones with any beds was 3.6, indicating that the extreme raw-count differential largely reflects population size, with a substantial residual per-capita gradient.

### 3.2 Road-network accessibility

Nationally, 90.1% of the routed population resided within 30 minutes’ drive of a facility with ICU beds, 97.8% within 60 minutes, and 99.2% within 90 minutes; 0.8% resided beyond 90 minutes. At the zone level, 190 of the 330 SMAs (87.9% of the routable population) had at least one facility with ICU-type beds within their own boundaries, and 140 did not (12.1% of the population; two remote island SMAs additionally could not be road-routed). Among residents of the 138 routable SMAs without in-zone ICU facilities, 40.8% could reach an out-of-zone ICU facility within 30 minutes and 84.7% within 60 minutes; 10.2% required 60–90 minutes and 5.1% more than 90 minutes. In 31 of these SMAs, however, a majority of residents were more than 60 minutes from the nearest ICU facility — and in 10 of them, effectively all residents were. Accessibility to the nearest ICU-or-HCU facility was higher throughout.

### 3.3 Nine-year evolution, 2016–2024

Total reported ICU/HCU beds grew from 16,136 in 2016 to 20,631 in 2024 (+27.9%), but the growth was asymmetric: HCU-type beds increased by 41.6% (9,548 to 13,517, peaking at 14,745 in 2021 before declining), whereas ICU-type beds increased by only 8.0% (6,588 to 7,114). The median SMA density rose from 10.4 to 12.6 per 100,000. The Gini coefficient of bed distribution declined from 0.384 to 0.262, falling in nearly every year. This equalization, however, was specific to the combined measure: computed on ICU-type beds alone, the fiscal year 2024 Gini coefficient was 0.365, with a median SMA-level ICU density of 2.8 per 100,000 population and a lower quartile of zero, reflecting the 140 SMAs without ICU-type beds. Throughout the period, the number of SMAs with zero ICU beds fluctuated only between 133 and 141 — effectively a frozen set of zones without intensive care capacity (Table 1).

**Table 1.**
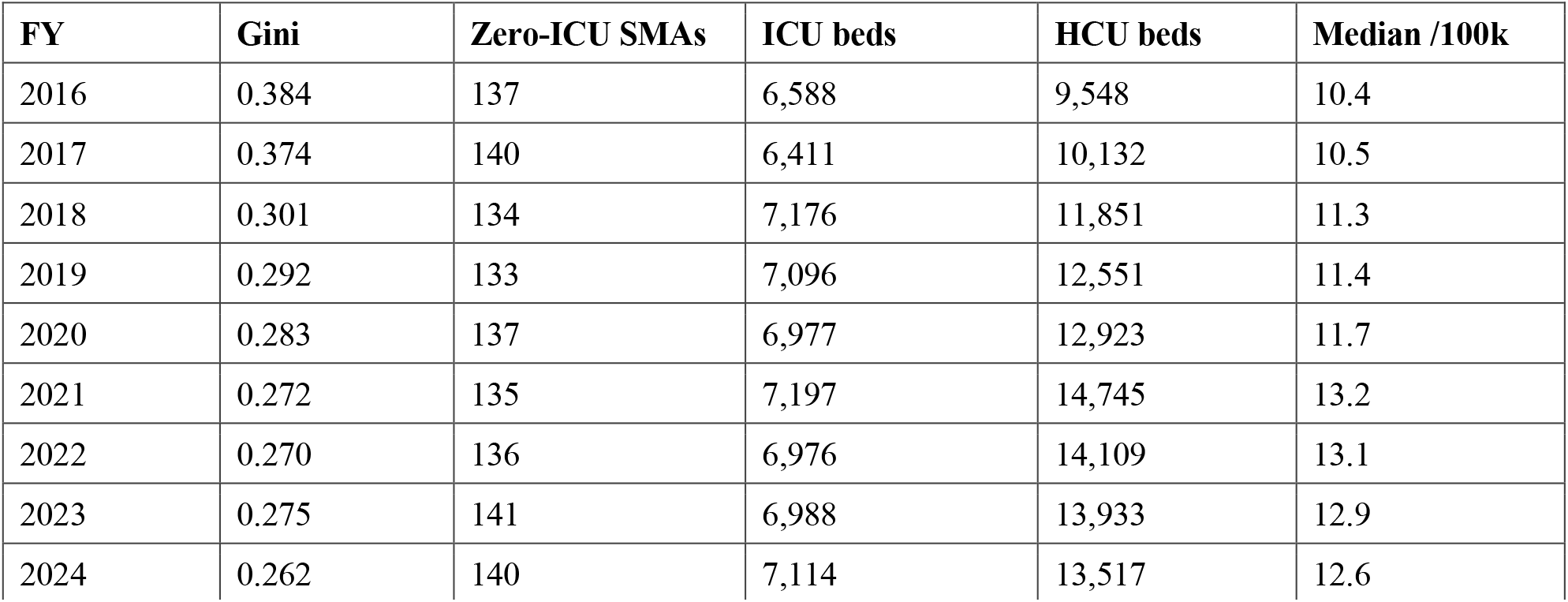
National trends in ICU/HCU supply across the 330 secondary medical areas of the 2025 classification, fiscal years 2016–2024. Gini coefficients are computed over SMA-level bed distribution; zero-ICU counts are based on reported ICU-type beds.

### 3.4 Structural persistence

Against these aggregate trends, the underlying supply structure was highly persistent. Of the 330 SMAs in the complete panel, 208 (63.0%) retained the same structural type across all nine years; 61 changed type once, 40 twice, and 21 three or more times. The 2016→2024 transition matrix (Table 2) shows that off-diagonal movement concentrated in exchanges between adjacent supply types, while the extremes — large multi-facility metropolitan configurations and low-density configurations — were nearly immobile. The supply-vulnerability ranking was likewise stable: Spearman correlations between adjacent years ranged from 0.887 to 0.924, and the correlation between 2016 and 2024 was 0.711, indicating strong preservation of the vulnerability hierarchy across nearly a decade.

**Table 2.**
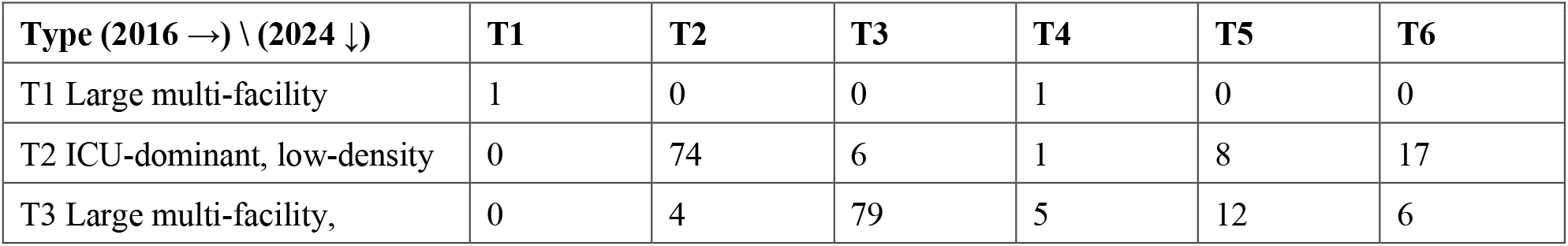
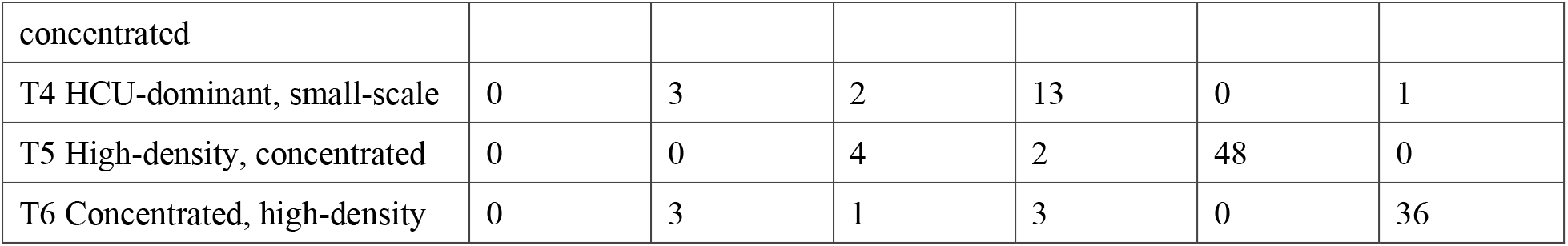
Transition matrix of structural types, 2016 → 2024, complete panel of 330 SMAs. Types were assigned in the 2024 anchor space (k-means, k = 6, on an 8-dimensional PCA basis); English type labels are working translations of the Japanese originals [TYPE LABELS TO CONFIRM WITH CO-AUTHORS].

### 3.5 The MeshScope-Region platform

All analyses are delivered through the MeshScope-Region web platform, which provides interactive SMA-level choropleths for each fiscal year, per-zone structural-type history strips, national trend panels, facility-level map markers, selectable inequality bases (combined ICU/HCU, ICU-only, and HCU-only Lorenz curves and Gini coefficients), and accessibility layers with selectable origins (zone centroids or population-weighted grid cells), refreshed annually from the underlying reports (Figure 5). Access to the platform environment can be provided by the corresponding author for replication and review purposes.

**Figure 1.**
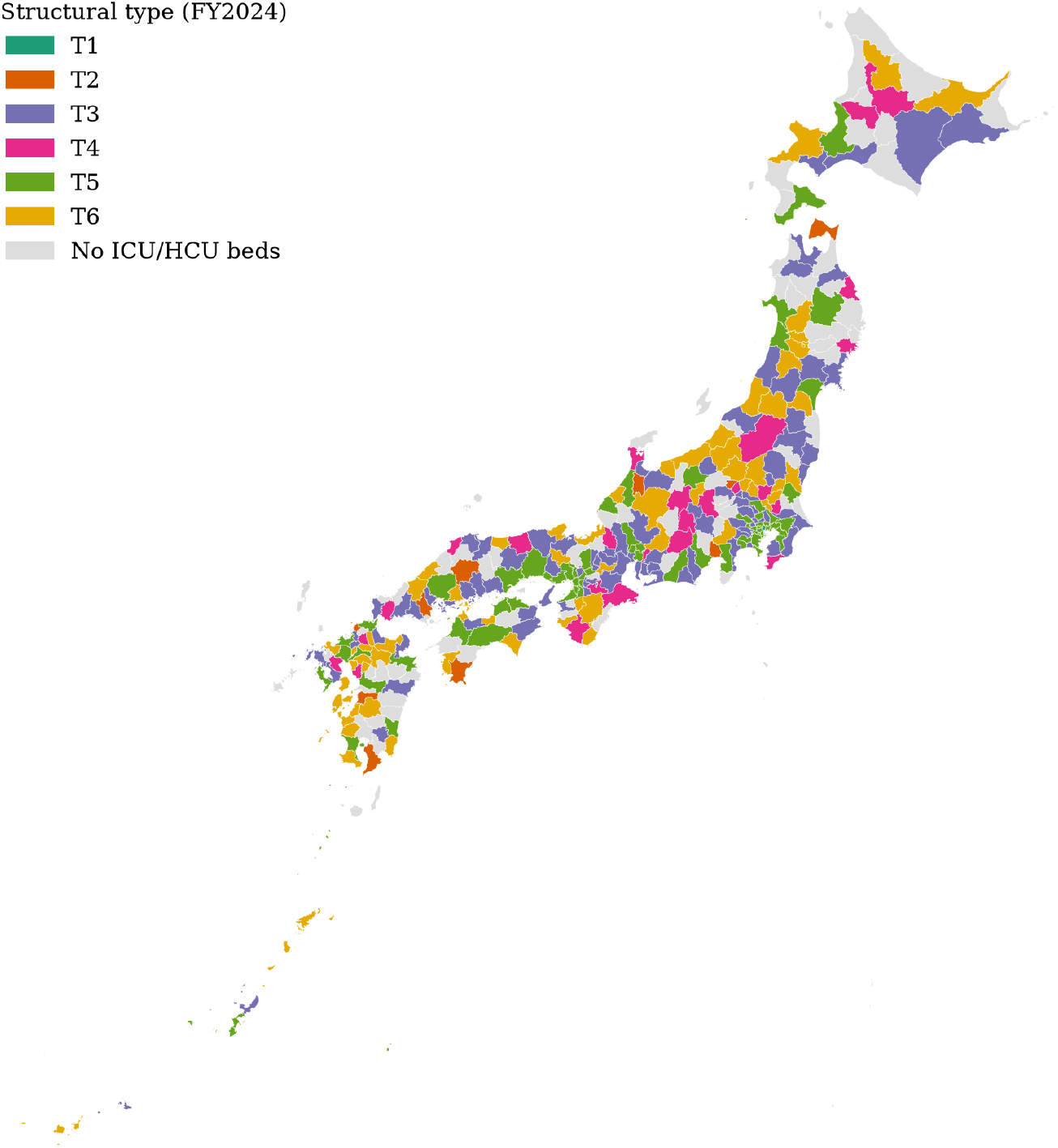
Choropleth map of structural types (T1–T6, k-means in the 2024 anchor space) across Japan’s 330 secondary medical areas (2025 classification), fiscal year 2024. Zones with zero reported ICU/HCU beds are shown in gray.

**Figure 2(a, b).**
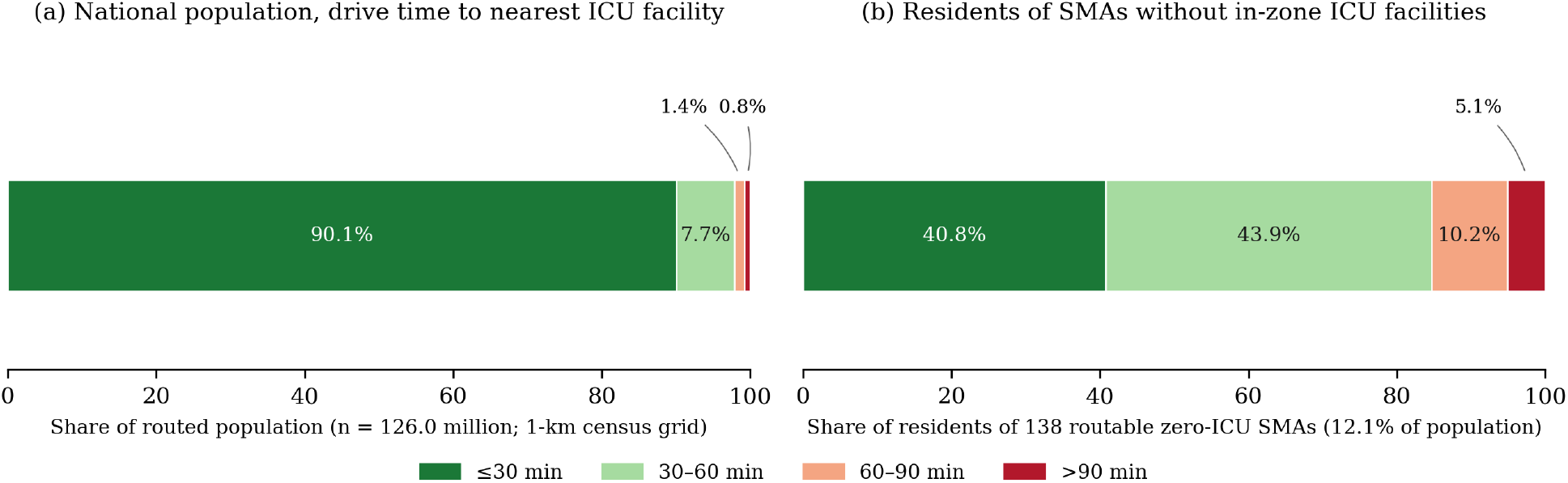
Drive-time accessibility to the nearest facility with ICU beds, computed with OSRM from 176,962 populated 1-km census grid cells. (a) National population coverage by drive-time band; (b) drive-time distribution for residents of the 138 routable SMAs without in-zone ICU facilities.

**Figure 2(c).**
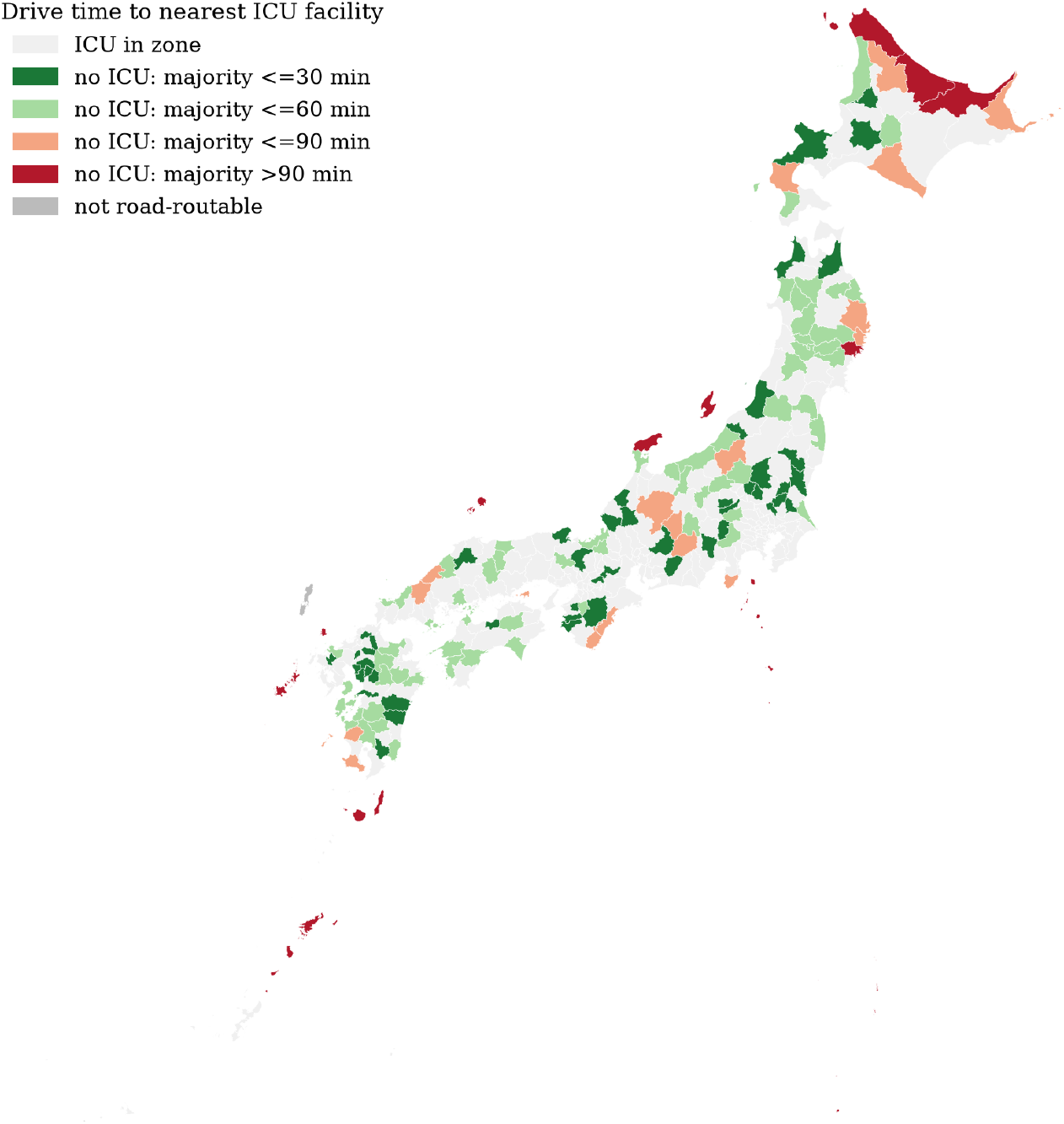
Map of SMAs by in-zone ICU availability; zones without in-zone ICU facilities are colored by the majority drive-time band of their residents to the nearest ICU facility, and island zones not reachable by road routing are shown in dark gray.

**Figure 3.**
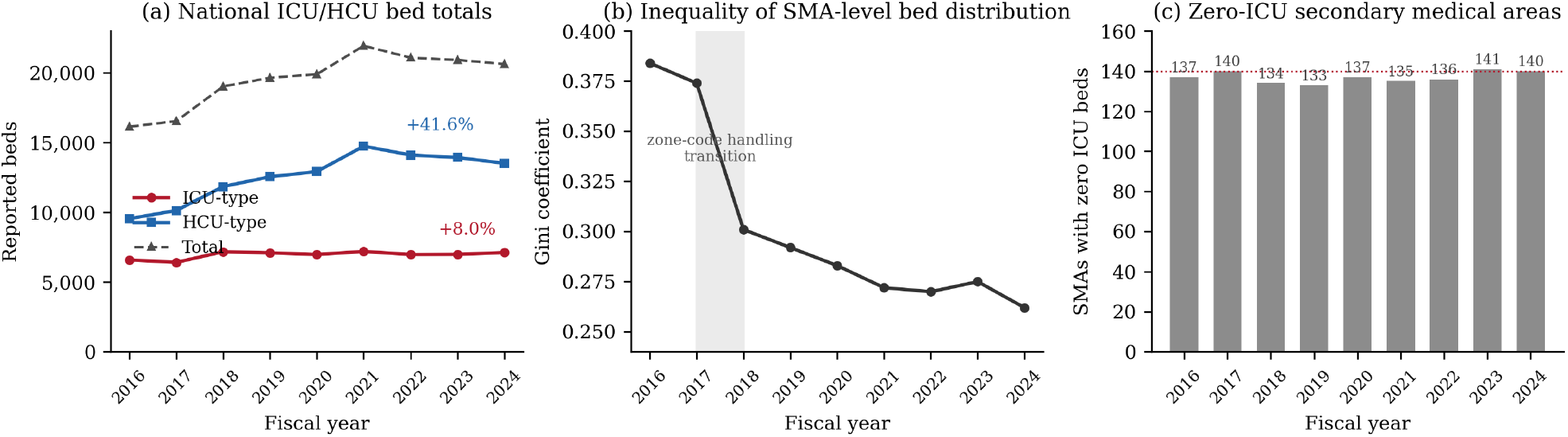
National trajectories, FY2016–2024: (a) ICU-type and HCU-type bed totals; (b) annual Gini coefficient of SMA-level bed distribution (shaded band: zone-code handling transition, see Limitations); (c) count of SMAs with zero ICU beds.

**Figure 4.**
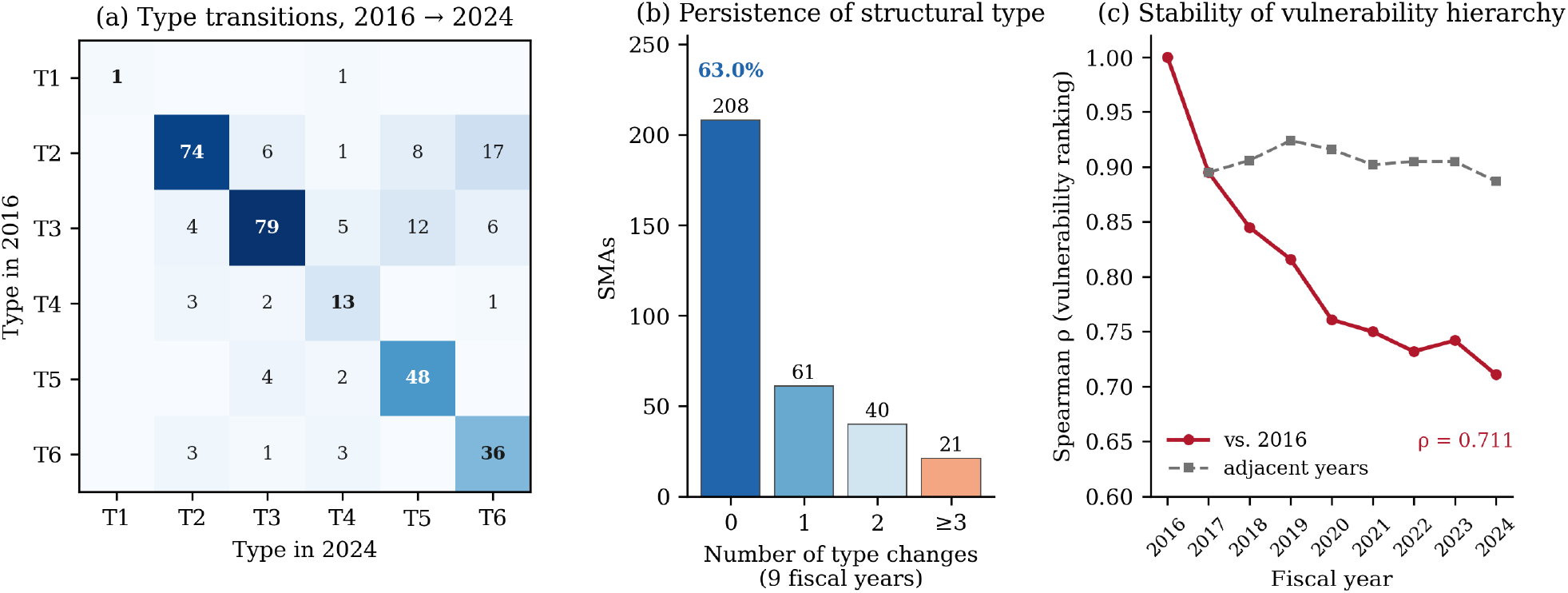
Structural persistence, FY2016–2024 (complete panel of 330 SMAs): (a) transition matrix of structural types between 2016 and 2024; (b) distribution of the number of type changes; (c) Spearman rank correlations of the supply-vulnerability index, against 2016 and between adjacent years.

**Figure 5.**
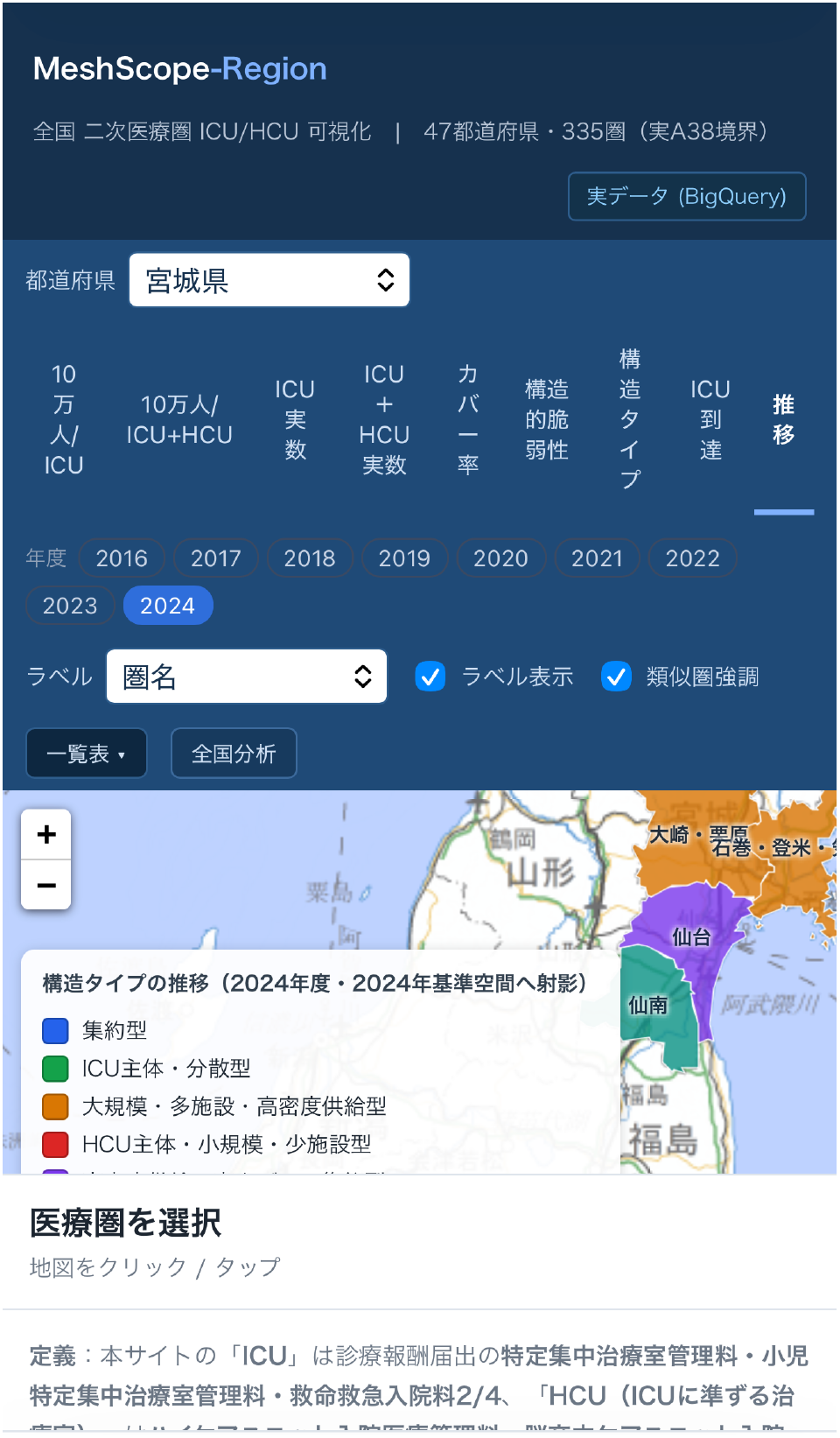
The MeshScope-Region platform (mobile view): the fiscal-year evolution tab with year chips (2016–2024), the structural-type choropleth (Miyagi Prefecture shown, FY2024, anchor-projected to the 2024 reference space), and the on-screen definition of ICU-type and HCU-type reimbursement categories corresponding to Section 2.1. [Internal note: the header zone count will be aligned to the 330-zone classification before journal submission.]

## 4. Discussion

### 4.1 Principal findings

Three findings stand out. First, ICU/HCU supply in Japan is structurally rigid: nearly two-thirds of SMAs did not change structural type in nine years, the supply-vulnerability hierarchy was strongly preserved (ρ = 0.711 across eight years), a stable set of approximately 140 SMAs has had no ICU beds throughout the period, and 74 SMAs report no critical care beds of any kind. Zone-level totals vary 229-fold in raw counts and 3.6-fold per capita at the 90th/10th percentile — an order of magnitude beyond the 8.6-fold prefecture-level variation previously reported [1]. Second, road-network accessibility substantially mitigates zone-level absence: although 140 SMAs lack an in-zone ICU facility, 84.7% of their residents are within 60 minutes’ drive of one, and only 0.8% of the national population is beyond 90 minutes. Third, the capacity growth of the past decade has been, in substance, an expansion of intermediate care: HCU beds grew 41.6% while ICU beds grew 8.0%, so aggregate bed counts increasingly overstate the growth of capacity for the most severe patients. Consistent with this, the fiscal year 2024 Gini coefficient computed on ICU-type beds alone (0.365) was markedly higher than the combined measure (0.262), indicating that part of the apparent equalization of critical care supply is an artifact of intermediate-care expansion.

### 4.2 Relation to prior work

Our results extend the prefecture-level evidence of Ohbe and colleagues [1, 2] by showing that the 8.6-fold prefecture-level variation conceals far larger heterogeneity at the SMA level, including a frozen set of zones with no ICU beds at all. They complement, and in one respect qualify, the recent SMA-level analysis by the same group [3]: whereas that study established that residents of SMAs without ICUs experience measurable disadvantages in access and outcomes, our accessibility layer shows that the population actually distant from intensive care is much smaller than the count of ICU-less zones suggests — the policy-relevant unit is drive-time coverage, not zone boundaries. Our contribution is complementary in design as well: reference [3] analyzes patient-level outcomes conditional on the supply map, whereas MeshScope-Region characterizes the supply map itself — its composition including HCU beds, its typology, its accessibility, and its nine-year dynamics — and publishes it as a standing platform. Compared with regional GIS analyses of emergency care accessibility [6], our accessibility layer is national, facility-complete for ICU/HCU, and refreshed alongside the underlying administrative data. Our structural typology likewise differs in its unit of analysis from facility-level classifications built on the same public data sources [8]: MeshScope-Region classifies planning areas rather than hospitals, and follows their types longitudinally.

### 4.3 Policy implications

The 8th Medical Plan cycle (2024–2029) and its successors require prefectures to justify critical care configurations within and across SMAs. Our findings suggest three concrete uses. First, the frozen set of zero-ICU SMAs should be triaged jointly with the accessibility layer: for the majority of such zones, whose residents reach an adjacent-zone ICU within 60 minutes, formalizing cross-zone referral and transport agreements may dominate bed construction; the 31 SMAs in which a majority of residents are more than 60 minutes from an ICU — together with the two island SMAs unreachable by road — are the candidates for targeted investment or air-transport arrangements. Second, the ICU-flat/HCU-growth pattern warrants explicit attention: if intermediate-care expansion substitutes for intensive-care capacity, surge resilience for the most severe patients will be overestimated by aggregate bed counts — a concern consistent with outcome evidence on intermediate care units operating without co-located ICUs [2]. Third, because the platform is refreshed annually from statutory reports, it can serve as a standing monitoring instrument for medical planning rather than a one-off survey. A companion probabilistic module (MeshScope-Scenario), performing seeded Monte Carlo capacity–demand simulation with variance-based sensitivity analysis grounded in the same audited dataset, extends this monitoring function toward prospective what-if analysis and will be described separately.

### 4.4 Strengths and limitations

The principal strengths are national completeness at the SMA level, the combination of capacity, composition, accessibility, and longitudinal structure in a single reproducible pipeline, and alignment with the statutory planning geography. Several limitations should be noted. First, reported bed counts reflect notified reimbursement categories, not staffed or operational beds; true surge capacity may differ, particularly under workforce constraints. Second, revisions to reimbursement categories over the study period enter the data as apparent structural change; the 2024 revision, for example, introduced new specific intensive care management fee subcategories. Third, all years were expressed in the 2025 SMA classification via a crosswalk spanning the 344-zone (2016–2017), 335-zone (2018–2023), and 330-zone (2024–) classifications, and zone codes missing in some report years (part of 2017 and 2018–2021) were imputed from the facility’s most recent observation. Ward-level bed totals evolve smoothly across fiscal 2017–2018, so the discontinuity in the Gini coefficient at that boundary (0.374 to 0.301) most plausibly reflects the transition in zone-code handling — from as-reported codes under the older classification to imputed current codes — rather than a reporting change in bed counts; year-over-year comparisons of inequality across that boundary should therefore be made cautiously. Fourth, the population denominator was fixed (2025 for the typology, 2020 census for accessibility), so per-capita trends reflect supply-side change only. Fifth, OSRM drive times assume free-flow conditions and do not model congestion, seasonal closures, or helicopter transport, and the zone-level accessibility layer summarizes each SMA by a representative point. Sixth, 1,432,234 residents in grid cells that could not be assigned to an SMA, and two remote island SMAs that could not be road-routed, are excluded from zone-level accessibility statistics (they are included in national mesh-level coverage where routable); because the unroutable zones are remote, their exclusion makes zone-level coverage estimates conservative in the favorable direction, and we report them separately. Finally, the structural typology depends on the anchor-space embedding [7]; alternative typologies could partition zones differently, although the persistence findings rest primarily on type stability and rank correlations, which are robust to relabeling.

### 4.5 Conclusions

MeshScope-Region integrates statutory reporting data into an SMA-level evidence base combining the distribution, composition, road-network accessibility, and nine-year evolution of Japan’s ICU/HCU capacity. The supply structure is regionally rigid and its growth has been concentrated in intermediate care, yet drive-time coverage of intensive care is far better than zone-level bed maps alone suggest. These findings, and the standing platform that produces them, are directly usable at the geographic unit at which Japanese health planning is conducted.

## Declarations

### Data availability

The SMA-level datasets and analysis payloads are available at [Zenodo/GitHub DOI]. Source data (Hospital Bed Function Reports, Population Census grid data) are publicly available from MHLW and e-Stat. The datasets and analytical structures generated during the current study are available from the corresponding author on reasonable request.

### Code availability

The data-integration and analysis code is available at [REPOSITORY].

### Author contributions

K.O.: conceptualization, methodology, software, data curation, formal analysis, visualization, writing — original draft. M.H.: data curation, validation, software. S.H.: supervision, domain expertise, writing — review and editing. All authors read and approved the final manuscript.

## Acknowledgements

Map data © OpenStreetMap contributors (ODbL). [OTHERS TO CONFIRM]

